# Comparison of Glomerular Filtration Rate Equations in a Rural New Mexico Cohort: Results from the COMPASS Study

**DOI:** 10.1101/2024.06.10.24308724

**Authors:** Monica Moya Balasch, Maria-Eleni Roumelioti, Christos P Argyropoulos

**Affiliations:** Division of Nephrology, Department of Internal Medicine, University of New Mexico School of Medicine, MSC 04-2785, Albuquerque, New Mexico, USA

**Keywords:** glomerular filtration rate equation, serum creatinine, cystatin C, beta-2-microglobulin, CKD-EPI, chronic kidney disease, diabetes, COMPASS, kidney function biomarkers

## Abstract

**Rationale and Objective:** The NKF-ASN Task Force recommends accurate kidney function estimation avoiding biases through racial adjustments. We explored the use of multiple kidney function biomarkers and hence estimated glomerular filtration rate (eGFR) equations to improve kidney function calculations in an ethnically diverse patient population.

**Study design:** Prospective community cohort study.

**Setting and Participants:** Rural New Mexico clinic with patients > 18 yo.

**Methods:** Markers of kidney function, IDMS-Creatinine (SCr), chemiluminescence Beta-2 Microglobulin (B2M), Nephelometry-calibrated ELISA Cystatin C (CysC), inflammation, glucose tolerance, demographics, BUN/UACR from the baseline visit of the COMPASS cohort, were analyzed by Kernel-based Virtual Machine learning methods.

**Results:** Among 205 participants, the mean age was 50.1, 62% were female, 54.1% Hispanic American and 30.2% Native American. Average kidney function biomarkers were: SCr 0.9 mg/dl, B2M 1.8 mg/L, and CysC 0.7 mg/dl. The highest agreement was observed between SCr and B2M-based eGFR equations [mean difference in eGFRs: (4.48 ml/min/1.73m^2^], and the lowest agreement between B2M and CysC-based eGFR equations (−24.75 ml/min/1.73m^2^). There was no pattern of association between the differences in eGFR measures and gender. In the continuous analyses, the absolute eGFR value (p<2 × 10^-16^) and serum albumin (p =6.4 × 10^-5^) predicted the difference between B2M- and SCr-based e-GFR. The absolute eGFR value (p<2 × 10^-16^) and age (p =7.6 x 10^-5^) predicted the difference between CysC- and SCr-based e-GFR.

**Limitations:** Relatively small sample size, elevated inflammatory state in majority of study participants and no inulin excretion rate measurements.

**Conclusion:** B2M should be strongly considered as a kidney function biomarker fulfilling the criteria for the NKF-ASN. B2M’s eGFR equation does not need adjustment for gender or race and showed the highest agreement with SCr-based eGFR equations.

## Introduction

Chronic kidney disease (CKD) is associated with various comorbid conditions and if left untreated can lead to end-stage kidney disease (ESKD) and renal replacement therapy^1,2^. Hence, the importance of accurate renal function estimation and renal health testing through the accurate calculation of estimated glomerular filtration rate (eGFR) is an ongoing topic of interest.

Appropriate GFR calculation can determine CKD prevalence and slow progression to ESKD^1^ through early detection of CKD and initiation of guideline appropriated therapy. However, GFR cannot be accurately measured in everyday practice and thus it is estimated using mostly serum creatinine (SCr) as a plasma filtration marker to predict kidney function^3,4^. Nevertheless, SCr levels can vary based on kidney tubular secretion, extrarenal elimination of SCr or generation by muscle or diet^5–7^.

Currently, eGFR calculations using the 2021 Chronic Kidney Disease Epidemiology Collaboration (CKD-EPI) equation adjust for gender and age ^4,8^. Prior to this adjustment, the eGFR equation was dependent on race ^9^ despite race being a social and not biological construct. Existing eGFR equations may not accurately calculate kidney function in diverse ethnic and racial populations. Studies have explored the potential of using emerging kidney filtration markers ^10,11^ such as serum Cystatin C (CysC) and serum Beta-2 Microglobulin (B2M) which may have less interpersonal variability based on muscle mass, age, and gender. However, there is no consensus on how these kidney filtration markers affect eGFR equations in indigenous and Hispanic populations. Thereby accurate CKD diagnosis and early therapeutic intervention may be delayed for these medically disadvantaged patient populations.

The state of New Mexico (NM) in the United States (US) is a disproportionately rural, low-income geographic region, with most of the population identifying as Native American and/or Hispanic American. Unsurprisingly, NM has one of the highest prevalence of diabetes which is one of the leading causes of ESKD in the US with over 60% of ESKD patients suffering from diabetes^12^. According to the American Diabetes Association approximately 200,548 people in NM (12.3% of the adults) have the diagnosis of diabetes with detrimental health-related and financial implications. In 2021, 3.9% of New Mexicans self-reported ever being told by a health professional that they have kidney disease compared to 3.0% in the general United States population^13^. Compounding on to the health disparities, these patient populations have been historically underrepresented in clinical research studies ^14–16^.

The National Kidney Foundation - American Society of Nephrology (NKF-ASN) Task Force recommends that kidney function be estimated by an approach that is accurate, in a manner that does not introduce bias through racial adjustments^17^. In order to fully understand how current eGFR equations that use SCr as a plasma filtration marker are a good predictor of kidney function compared to equations that use non-creatinine markers, we conducted a sub-analysis from the COMPASS study in rural northern NM ^12^. In the COMPASS study population with the majority identifying as Hispanic and Native American and in a region with extremely high prevalence of glucose intolerance and diabetes, we hypothesized that the various eGFR equations that use different serum biomarkers (serum SCr, CysC, or B2M) will evaluate kidney function differently.

## Methods

### Settings and Study subjects

This is a sub-analysis of a cross-sectional study involving participants who lived in or within 20 miles from an apparent hotspot of CKD in rural New Mexico who were between 18 - 80 years old and were able to sign an informed consent form. Study methods and design are documented in the design paper^12^; briefly, eligible participants did not receive chronic dialysis treatment or had not received a functioning kidney transplant in the past. We identified 218 adult participants with these characteristics from March 2016 through February 2020 and examined the data from their first study visit for the COMPASS study^12^.

The study protocol was reviewed and approved by the University of New Mexico Health Sciences Center, Human Research Protection Office on December 4th, 2015 under Study ID: 15–575.

### Sample and Data collection

Once the informed consent was signed, participants underwent a laboratory evaluation of non-fasting blood and urine samples in addition to the collection of medical history items and a physical exam. Patients were classified as having a positive CKD screen based on impaired eGFR (CKD G3–5) and/or albuminuria (CKD A2–3)^18^

The medical history questionnaire incorporates elements from previous community screening efforts for CKD (e.g., NKF, Kidney disease Early Evaluation Program, KEEP^19,20^) and national health surveys (NHANES) ^21,22^. Self-reported demographics (age, gender, race, ethnicity), social history, family history, list of current medications, attained education level and health insurance provider(s) were also collected.

Physical examination consisted of measurement of systolic and diastolic blood pressure (SBP and DBP) in both arms using appropriately sized cuffs, recording of height and weight for BMI calculation, number of respirations and blood oxygen saturation.

### Clinical Laboratory Measures

Extensive laboratory testing was performed on the non-fasting plasma and urine samples of study participants. A cold chain was set up for the transport of specimens collected at the study site to the UNM Clinical and Translational Science Center (CTSC) laboratory that performed the laboratory assessments. Clinical laboratory measures, sample aliquots and stores for further testing were handled by the CSTC laboratory and the regional reference lab Tricore.

The specimens collected produced results for serum glucose levels, hemoglobin A1C (HbA1c), complete blood cell count (hemoglobin and hematocrit), a full metabolic panel, high sensitivity C-reactive protein (hs-CRP), kidney function panel (SCr, blood urea nitrogen, albumin), serum and urine osmolality, urine albumin to creatinine ratio (UACR), urinalysis and urine culture, intact parathyroid hormone (iPTH), Vitamin D levels, urine electrolytes (phosphorus and calcium), urine uric acid, serum B2M and serum CysC. Serum B2M was measured at the Tricore laboratories by immunonephelometry.

Serum CysC was measured with a low cost Luminex research-grade assay (R&D systems), which is more economical than a reference immuno-nephelometry assay used by clinical laboratories. Serum CysC was measured with both techniques in a calibration sub-study involving 20 individuals; the latter provided linear calibration factors to convert the Luminex assay readings to immuno-nephelometry ones via the formula:

Immunonephelometric CysC (mg/L) = 0.01 x (ELISA cystatin in ng/ml) - 0.065

### Estimated Insulin Sensitivity

Quantitative Insulin Sensitivity Check Index (QUICKI: 1/(ln(insulin)+ln(glucose)) and Homeostasis Model Assessment of Insulin Resistance (HOMA-IR) calculations were also provided due to non-fasting blood samples collected. QUICKI and HOMA-IR are surrogate indexes for insulin sensitivity/resistance derived from blood insulin and glucose concentrations when the patient is fasting (steady condition) or after an oral glucose load (dynamic conditions). Specifically, QUICKI is an empirically derived mathematical transformation of fasting blood glucose and plasma insulin concentrations that provides a simple, robust, accurate and reproducible method that can predict insulin sensitivity changes after the onset of diabetes or therapeutic interventions. HOMA is a model of interactions between glucose and insulin concentrations for a wide range of combinations of insulin resistance and β-cell function. Most studies using HOMA use an approximation through a simple equation to derive a surrogate index of insulin resistance^23^. Previous studies have shown that fasting and non-fasting values correlate closely and are sufficient for population-based research where it is difficult to ensure study subjects are fasting^24^.

### Estimated Glomerular Filtration Rate

We also aimed to study the potential equation adjustments for gender and age. GFR (mL/min/1.73m^2^) was estimated using the 2021 CKD-EPI Creatinine, 2012 CKD-EPI Cystatin C, CKD-EPI Cystatin-Creatinine, and CKD-EPI Beta-2 Microglobulin equations. These equations are detailed in the supplement and are free of racial adjustments.

CKD was defined as either reduced renal function (low eGFR) and or albuminuria >30 mg/g creatinine. Albuminuria was considered any value above 30 mg/g creatinine on Albumin-to-Creatinine Ratio in a spot urine sample. e-GFR values <60 mL/min/1.73 m^2^ were considered to represent reduced kidney function.

### Statistical analysis

Patient population demographics and laboratory values were summarized as mean, percentage (%) and standard deviation (SD). The agreement between the four eGFR equations was tested using Bland-Altman plots. An exploratory data analysis with multivariate clustering techniques was undertaken to better visualize how well eGFR estimates from the three single marker equations track each other. Clustering resolved the eGFRs according to whether they followed the same direction (all increasing or all decreasing, i.e. concordant) vs. those exhibiting discordant behavior (i.e. two of the markers pointing in the same direction, while the last one did not).

Finally, we explored whether the discordancy could be explained via demographics, systolic and diastolic blood pressure measurements, levels of markers of inflammation, glucose intolerance, albuminuria, and other laboratory values (e.g. BUN and albumin that were incorporated in the first estimating equation for eGFR, i.e. the 4 variable MDRD formula) by null-space Kernel regression-based learning methods^25^.

Advanced modeling techniques allow the effect of predictors to be flexibly estimated from the data via thin plate penalized splines or shrink the estimated degrees of freedom of the relationship to zero, effectively eliminating these predictors from the model. For our project we built two such models: a binary one which attempted to explain being assigned to the discordant cluster, and a continuous one in which each of the two non SCr-based eGFR equations were related to the 2021 CKD-EPI SCr-based eGFR equation along with the other predictors. The former model captures the semi qualitative discordance identified by the clustering models, while the latter ones utilize the continuous nature of the eGFR predictions to explain numerical deviations of the CysC and B2M CKD-EPI equations from the SCr-based CKD-EPI equation. All analyses were done in R language (v 4.2, packages cluster and mgcv). Given the exploratory nature of these analyses, we set the level of statistical significance to the more conservative value of 0.01 rather than 0.05 to minimize the potential for false positive findings.

## Results

### Subject Characteristics

For this report we utilized data from 218 study subjects enrolled in the COMPASS study and 205 had complete data available for analysis (**Figure 1, STROBE Diagram**). **Table 1** summarizes the demographic and anthropometric characteristics of the study population along with their SBP and DBP measurements that occurred during Visit 1 of the COMPASS study. **Table 2** summarizes basic laboratory values including serum and urine-based kidney function markers, insulin sensitivity markers, and the eGFR estimates using the equations detailed in the methods section.

**Figure 1:**
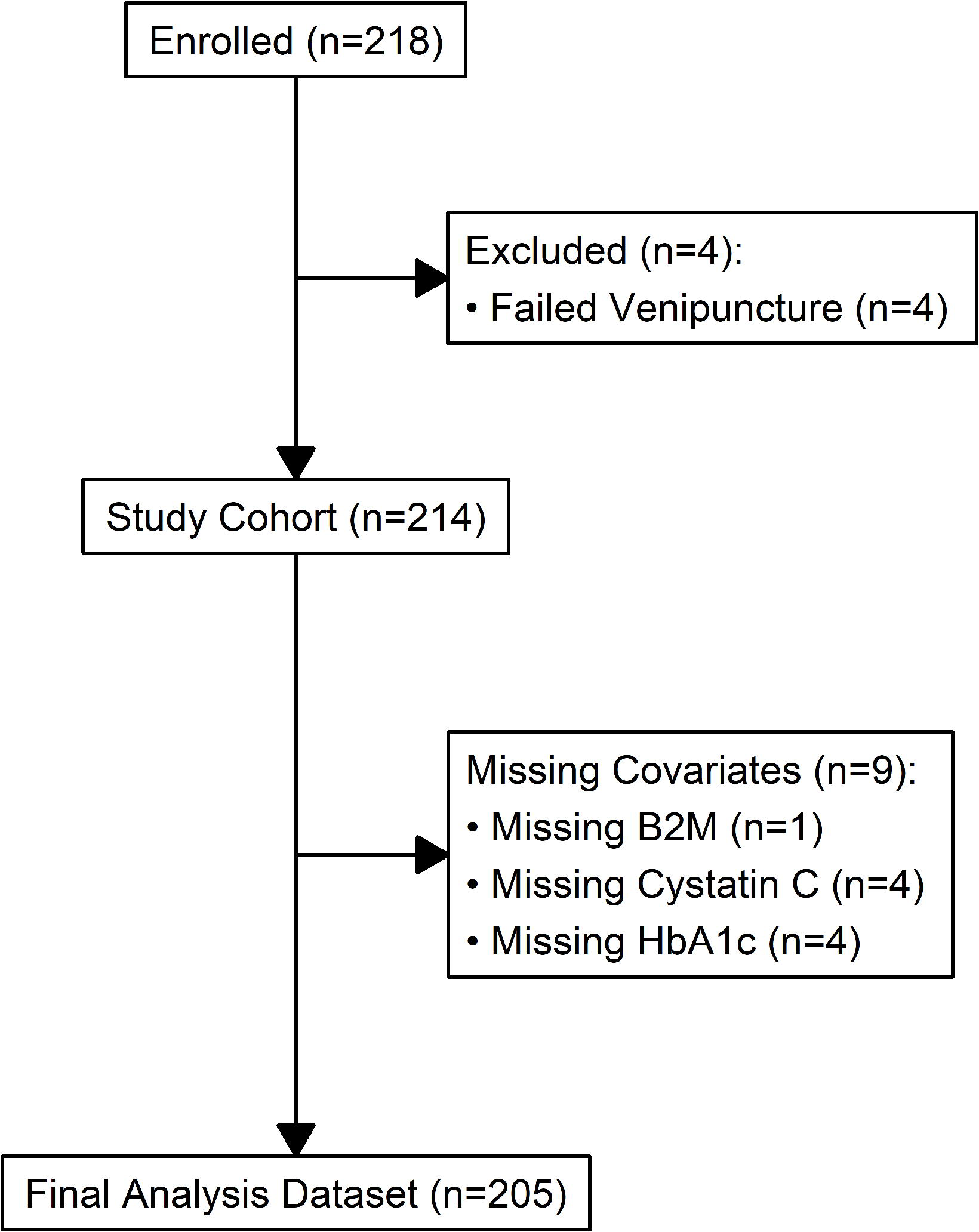
STROBE diagram for the COMPASS cohort.

**Table 1.**
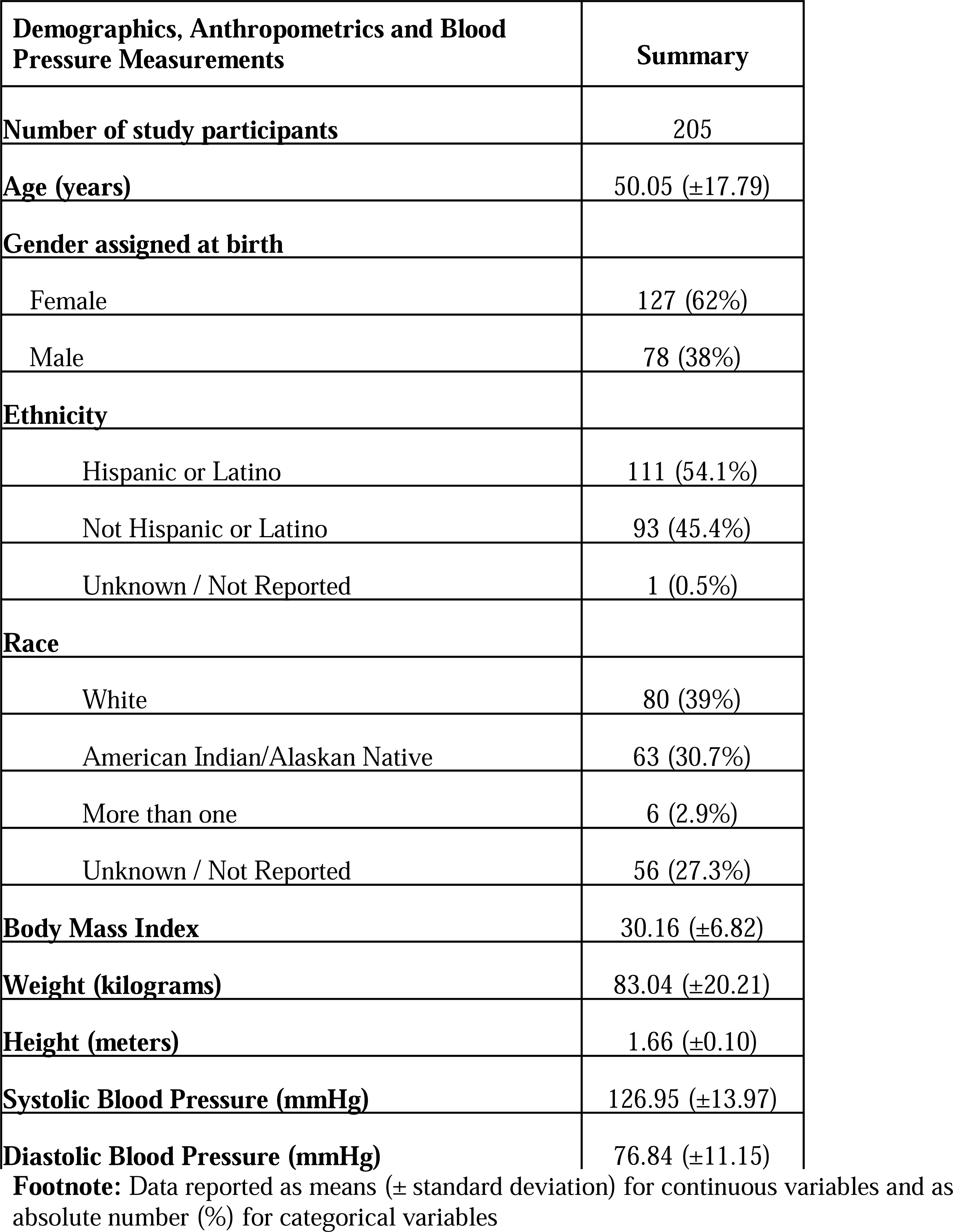
Cohort characteristics.

| **Demographics, Anthropometrics and Blood Pressure Measurements** | **Summary** |
| --- | --- |
| **Number of study participants** | 205 |
| **Age (years)** | 50.05 (±17.79) |
| **Gender assigned at birth** |  |
| Female | 127 (62%) |
| Male | 78 (38%) |
| **Ethnicity** |  |
| Hispanic or Latino | 111 (54.1%) |
| Not Hispanic or Latino | 93 (45.4%) |
| Unknown / Not Reported | 1 (0.5%) |
| **Race** |  |
| White | 80 (39%) |
| American Indian/Alaskan Native | 63 (30.7%) |
| More than one | 6 (2.9%) |
| Unknown / Not Reported | 56 (27.3%) |
| **Body Mass Index** | 30.16 (±6.82) |
| **Weight (kilograms)** | 83.04 (±20.21) |
| **Height (meters)** | 1.66 (±0.10) |
| **Systolic Blood Pressure (mmHg)** | 126.95 (±13.97) |
| **Diastolic Blood Pressure (mmHg)** | 76.84 (±11.15) |

**Footnote:** Data reported as means ( ± standard deviation) for continuous variables and as absolute number (%) for categorical variables

**TABLE 2.**
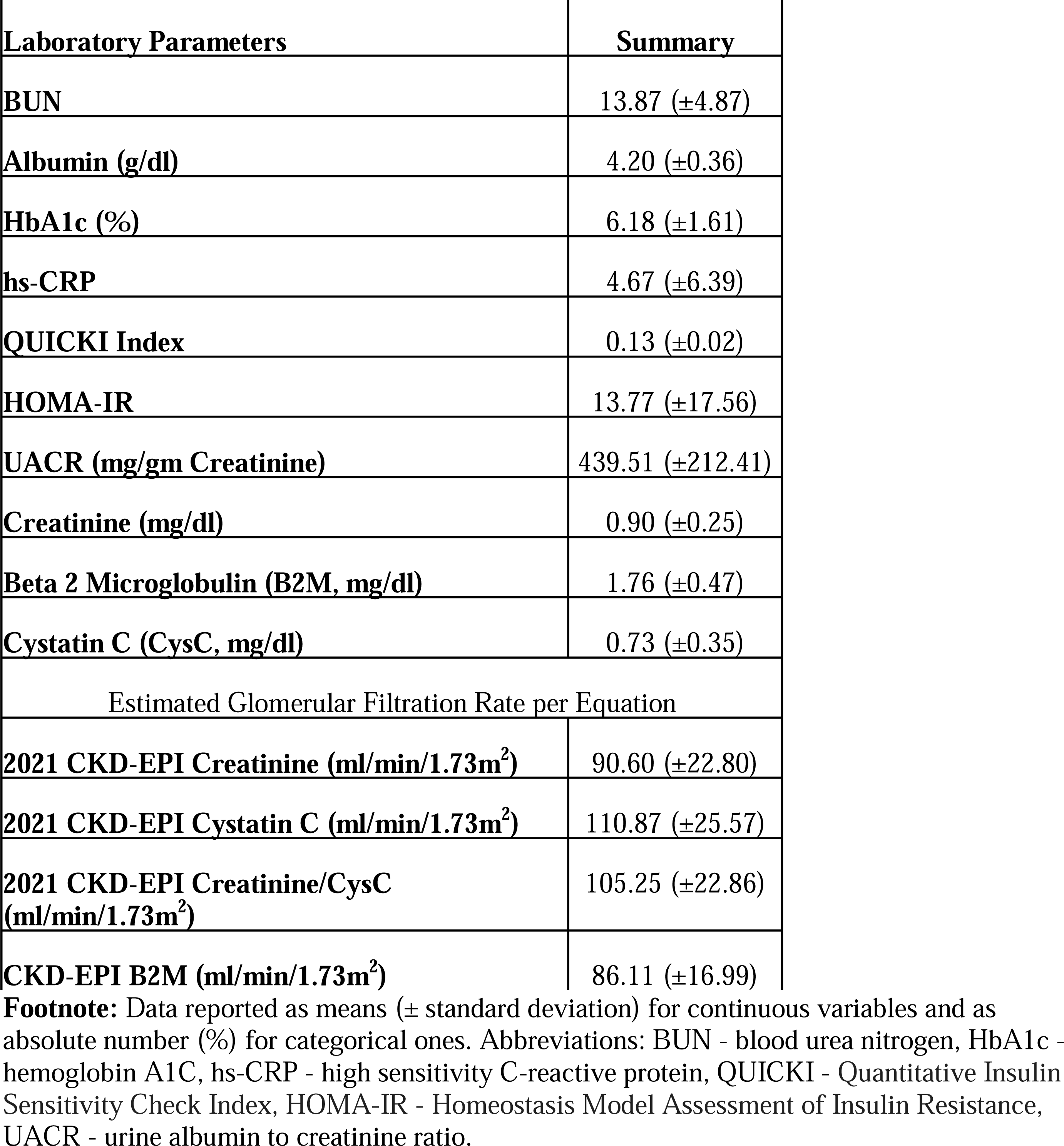
Laboratory values.

| **Laboratory Parameters** | **Summary** |
| --- | --- |
| **BUN** | 13.87 ($\pm 4.87$) |
| **Albumin (g/dl)** | 4.20 ($\pm 0.36$) |
| **HbA1c (%)** | 6.18 ($\pm 1.61$) |
| **hs-CRP** | 4.67 ($\pm 6.39$) |
| **QUICKI Index** | 0.13 ($\pm 0.02$) |
| **HOMA-IR** | 13.77 ($\pm 17.56$) |
| **UACR (mg/gm Creatinine)** | 439.51 ($\pm 212.41$) |
| **Creatinine (mg/dl)** | 0.90 ($\pm 0.25$) |
| **Beta 2 Microglobulin (B2M, mg/dl)** | 1.76 ($\pm 0.47$) |
| **Cystatin C (CysC, mg/dl)** | 0.73 ($\pm 0.35$) |
| **Estimated Glomerular Filtration Rate per Equation** |  |
| **2021 CKD-EPI Creatinine (ml/min/1.73m<sup>2</sup>)** | 90.60 ($\pm 22.80$) |
| **2021 CKD-EPI Cystatin C (ml/min/1.73m<sup>2</sup>)** | 110.87 ($\pm 25.57$) |
| **2021 CKD-EPI Creatinine/CysC (ml/min/1.73m<sup>2</sup>)** | 105.25 ($\pm 22.86$) |
| **CKD-EPI B2M (ml/min/1.73m<sup>2</sup>)** | 86.11 ($\pm 16.99$) |

**Footnote:** Data reported as means ( ± standard deviation) for continuous variables and as absolute number (%) for categorical ones. Abbreviations: BUN - blood urea nitrogen, HbA1c - hemoglobin A1C, hs-CRP - high sensitivity C-reactive protein, QUICKI - Quantitative Insulin Sensitivity Check Index, HOMA-IR - Homeostasis Model Assessment of Insulin Resistance, UACR - urine albumin to creatinine ratio.

The mean age of all study participants was 50.1 (± 17.8) years. Study participants reflected the community population and were racially diverse (American Indians approximately 30%), but predominantly self-reported to be white (approximately 39.0%) and Hispanic (54.1%). The majority were women (62%), and the average BMI was in the overweight range (30.2 ± 6.8).

### Limits of Agreement and Machine Learning Analysis of eGFR measures

The highest agreement was observed between SCr and B2M-based eGFR equations (mean difference in eGFRs: 4.48, 95% limits of agreement −33.76 to 42.73 ml/min/1.73m^2^), followed closely by the agreement between the 2012 Cystatin C eGFR equation and the 2021 combined SCr and CysC eGFR equations (mean difference in eGFRs: 5.62, 95% limits of agreement - 13.90 to 25.23 ml/min/1.73m^2^). However, the agreement between the 2021 SCr and the combined SCr/CysC equations was considerably less: mean difference of −14.65 (95% limits of agreement −42.36 to 13.05 ml/min/1.73m^2^).

The mean difference between the 2021 SCr and the 2012 CysC eGFR equations was −20.27 (95% limits of agreement −65.45 to 24.95 ml/min/1.73m^2^). The greatest lack of agreement was observed between B2M and CysC-based eGFR equations (mean difference −24.75, 95% limits of agreement −69.26 to 19.75 ml/min/1.73m^2^) (**Figure 2**). There was no obvious pattern of association between the differences in eGFR measures and gender.

a. Clustering Analysis

**Figure 2.**
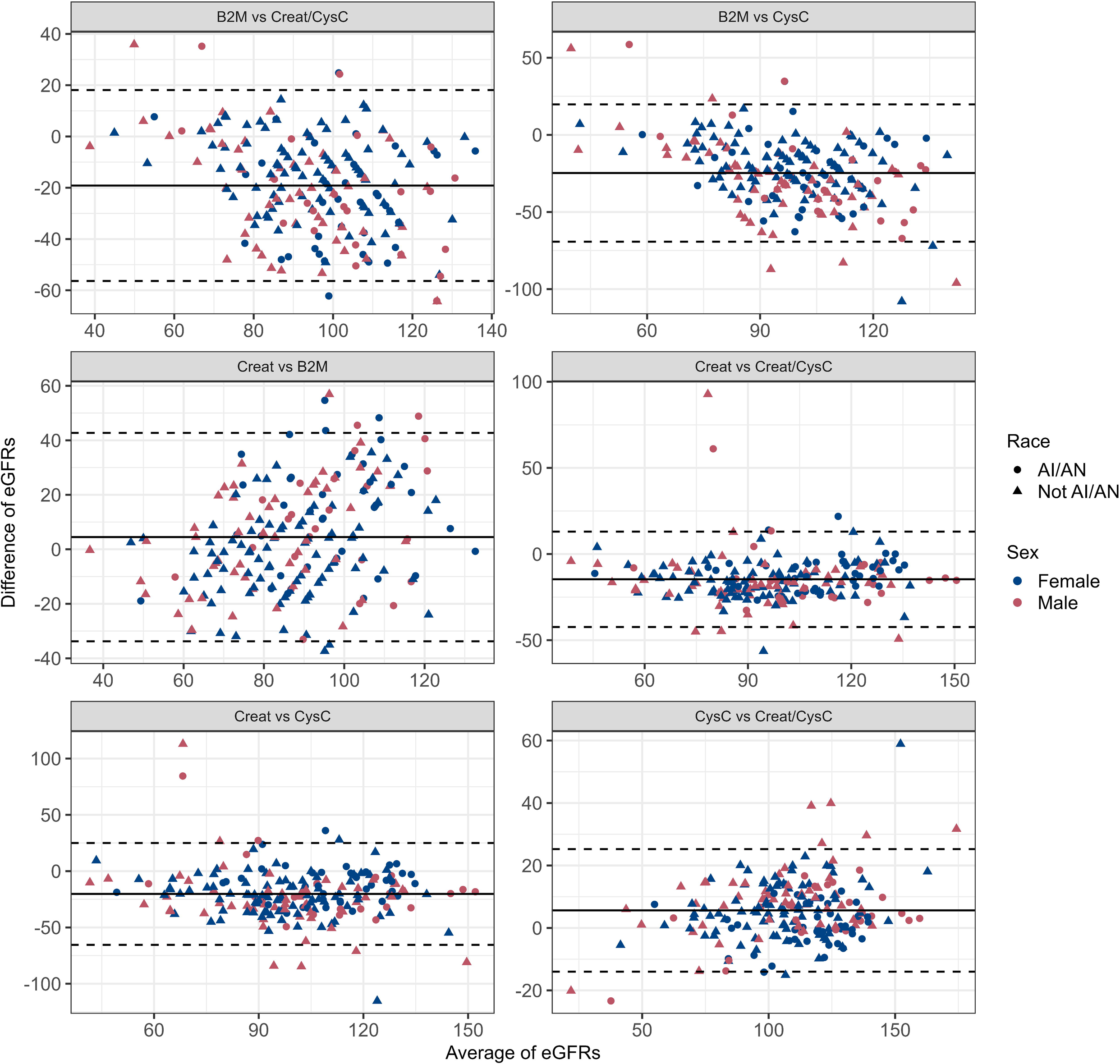
Bland Altman Analysis for the eGFR measures.

The clustering analysis showed that most measurements were concordant and only 11% were discordant (**Figure 3**). None of the factors of ethnicity, age, gender, glycemic control (HbA1c, HOMA-IR, QUICKI), albuminuria, SBP and DBP, height, weight, serum albumin or serum BUN could predict the discordant status of the eGFR measurements (not shown, all p-values > 0.11).

1. b) Continuous Analyses

**Figure 3.**
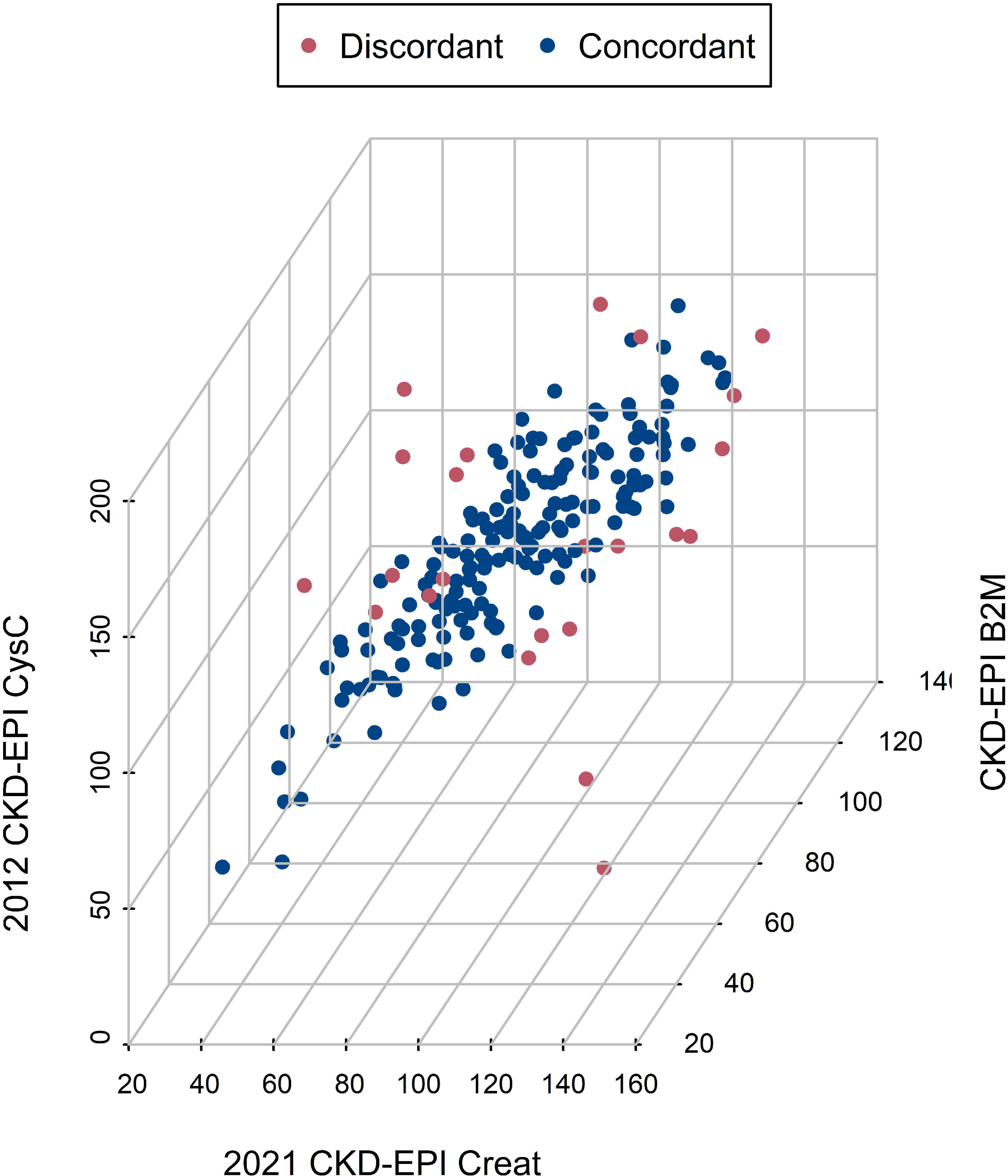
Machine Learning Clustering Analysis of the three-single marker eGFR equations.

In the continuous analyses we identified the following predictors of the difference between the eGFR of B2M and SCr-based equations: the absolute value of eGFR (p < 2 x 10^-16^) and the level of serum albumin (p =6.4 x 10^-5^). Predictors of the difference between the eGFR of CysC and SCr-based equations: the absolute value of the eGFR (p < 2 x 10^-16^) and age (p =7.6 x 10^-5^). The model for the difference between B2M and SCr eGFR equations accounted for a higher percentage of the variance than the model for the combined SCr and CysC eGFR equations (58.1% vs 29.5%).

1. c) Kernel Methods

The Kernel methods estimated relations are shown in **Figure 4**. Either B2M or CysC eGFR equation estimates were lower than the SCr eGFR equation estimates when the latter’s value was in the hyperfiltration range (>100ml/min/1.73m^2^) and higher in the range between 40 to 60 ml/min/1.73m^2^. CKD-EPI B2M-based eGFR was smaller than the CKD-EPI SCr eGFR outside the “normal” range of albumin (4-4.6 g/dl), while age outside the span of 40-60 years old affected the relationship between CKD-EPI CysC and CKD-EPI SCr.

**Figure 4.**
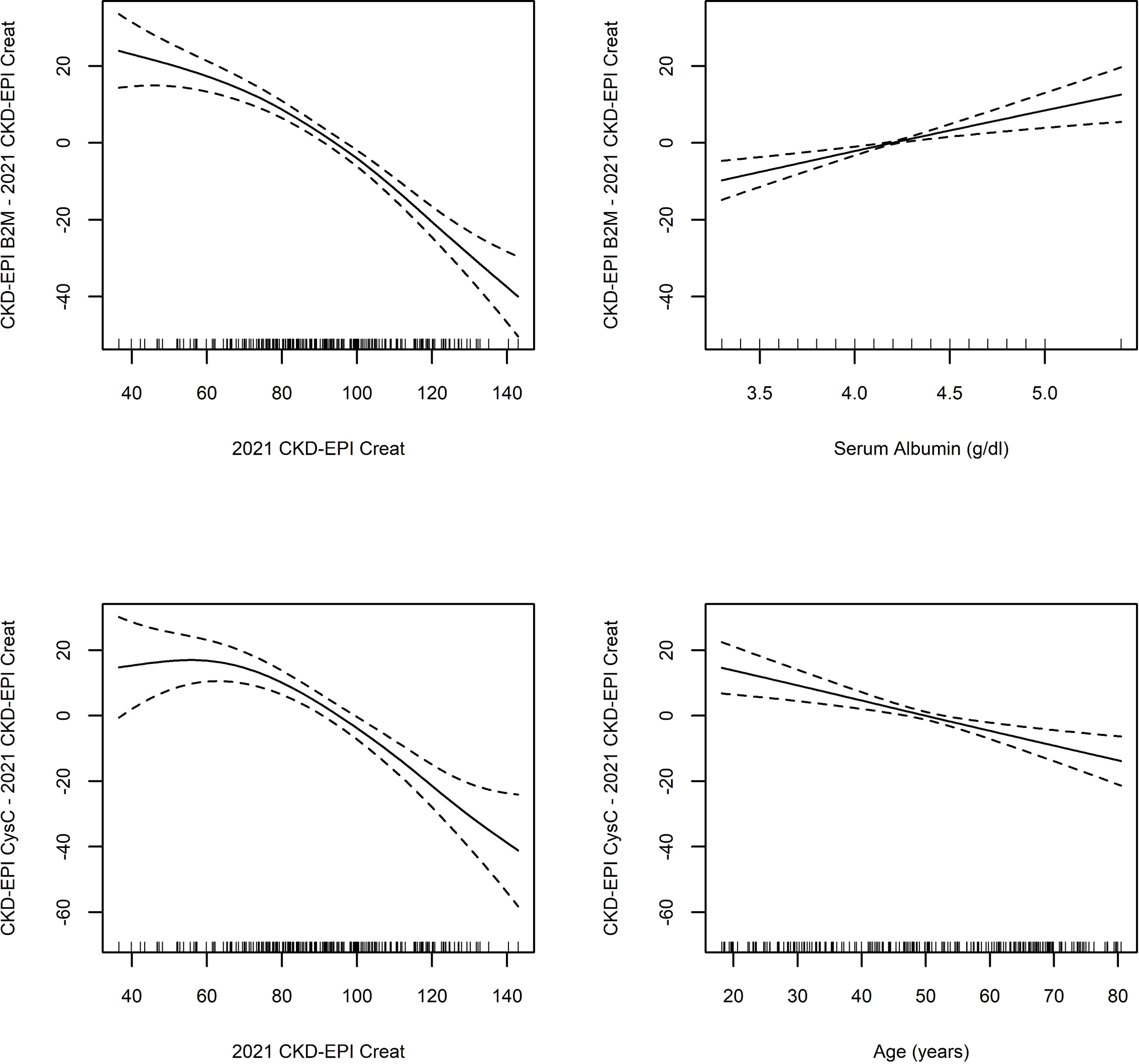
Correlates of differences between B2M and Creatinine eGFR; and Cystatin and Creatinine.

## Discussion

Existing eGFR equations that use SCr as a kidney function biomarker and adjust for age and gender may not accurately estimate kidney function in diverse ethnic and racial populations. To our knowledge our study is the first to explore the agreement between among four different eGFR equations incorporating those filtration markers in an underrepresented diverse rural patient population (majority of study participants identified as Hispanic or Native American).

After completion of our analyses the highest agreement was observed between SCr and B2M-based eGFR equations, and the lowest agreement between B2M and CysC-based eGFR equations. When we examined the three biomarkers of kidney function (creatinine, cystatin C and beta 2 Microglobulin) in this unique population, concordance was noted for 75%-83% of the biomarkers when their raw measurements and eGFR estimates were analyzed via clustering analyses, while no clinical factor was predictive of the discordance.

In this report the absolute eGFR value and age predicted the difference between CysC- and SCr-based eGFR. CysC is an endogenous low-molecular weight protein used in the estimation of kidney filtration. In the 2012 guidelines by the Kidney Disease Improving Global Outcomes (KDIGO) 1 CysC was recommended as a CKD confirmatory test^26,27^ and is deemed helpful in conditions affecting SCr levels such as extremes of muscle mass, severe chronic illness, and advanced age ^27,28,29^. However, inflammation, steroid treatment and thyroid disease are also well-known non-GFR determinants of serum CysC^27,30^.

A large national study (CRIC) of 1248 adults with CKD conducted cross-sectional analyses of baseline data such as self-reported race, genetic ancestry markers, and SCr, serum CysC, and 24-hour urinary Cr levels. They found that the precision and validity of eGFR from serum CysC, a renal filtration marker currently available in the lab, were similar to those of eGFR measurements based on the SCr levels, without having to consider race or ancestry^8^. Other studies with diverse patient populations that have looked at estimation of GFR with CysC^31,32^, without taking into consideration race, have also found that this may be a better biomarker.

Discordances between eGFRs derived from Scr and CysC are common and predict serious adverse outcomes. Recently, Carrero et al., 2023 published their results from an observational study on 158,601 adults (48% women; mean age 62 years, eGFRcr 80, and eGFRcys 73 mL/min/1.73/m^2^) undergoing testing for creatinine and CysC on the same day in connection with a health care encounter during 2010-2018 in Stockholm, Sweden^33^. In this study, they found that discordances between eGFRcys and eGFRcr are common, and 1 in 4 patients tested had an eGFRcys > 28% lower than their eGFRcr. They also showed that when eGFRcys was lower than the eGFRcr it consistently identified patients at higher risk of adverse outcomes such as cardiovascular events, kidney replacement therapy, acute kidney injury, and death. Within the same year Farrington et al., 2023 published results from a study that sought to enhance the knowledge of the risk factors and clinical implications of having a large eGFR discrepancy^27^.

Participants in the Atherosclerosis Risk in Communities Study^34^, a prospective cohort study of US adults, were followed over 25 years. eGFR discrepancy was measured at five clinical visits and defined as eGFRcys either 30% lower or higher than eGFRcr, the current clinical standard of care. Having eGFRcys lower than eGFRcr was associated with worse kidney-related laboratory derangements and a higher risk of adverse health outcomes as well.

Non kidney related factors such as ethnicity, inflammation and diabetes are candidates for factors that may explain the discordance among the various known kidney biomarkers. Clinically variable adjustments only marginally improve agreement among kidney function serum biomarkers. A study compared eGFRCr and eGFRCysC trends among 1069 patients from the Korean CKD cohort (KNOW-CKD), which enrolls predialytic CKD patients, whose creatinine and CysC had been followed for more than 4 years^35^. They found that young age and proteinuria were related to discrepancies in trends between the two eGFRs. In their discussion they argue that this is because most filtered CysC is reabsorbed and metabolized by the proximal tubule cells. Another study by Liu X et al (AJKD 2016) identified that the concentration of CysC was influenced by the urine protein excretion, an influence stronger than that of SCr^36^. Several other studies suggest that heavy proteinuria influenced renal handling of CysC^37,38^. A previous analysis of the discordance along low molecular weight filtration markers (e.g. cystatin C, beta 2 Microglobulin and beta trace protein) among community dwelling elderly adults showed that these protein biomarkers are affected less by age and sex and not affected by ethnicity compared to creatinine^5^. Our clustering and kernel-based analyses confirm these findings in a population of Hispanic and Native Americans.

As the replacement of race-adjusted SCr-eGFR with CysC-eGFR gains momentum, it is important to understand potential analytical biases introduced by CysC in other racial and ethnic groups.

In our study, B2M was affected the least by race and ethnicity. This should be strongly considered as a measure fulfilling the criteria for the NKF-ASN organizations because its eGFR equation does not need adjustment for age, race or gender and thus represents a more optimal method to calculate eGFR. Our observations suggest that B2M or CysC can be used to obtain an estimate of eGFR when the latter is in the hyperfiltration range, thus completing the suggested use of these alternative biomarkers when eGFR creatinine is in the 40-60 mL/min/1.73 m^2^ range and CKD diagnosis confirmation is needed studies^39^.

In the continuous analyses of our report the absolute eGFR value and serum albumin predicted the difference between B2M- and SCr-based eGFR. The effect of serum albumin on the difference between B2M and Creatinine based formulas of eGFR may be related to the biological function of B2M. The neonatal Fc receptor present in all cells is a B2M associated protein responsible for recycling albumin^40^. Our patient population demonstrated an inflammatory state with elevated hs-CRP. In these high turnover states, B2M shedding may be occurring which could simultaneously lower albumin and B2M based eGFR. However, we were not able to demonstrate a relationship between the hs-CRP and the differences of B2M – and SCr-based EPI equations derived eGFRs, likely because the albumin may be a more sensitive marker of such processes.

Our data suggest a potentially optimal way to select eGFR protein biomarkers when a confirmatory measurement of a creatinine based eGFR is needed. This approach would incorporate the patient’s, age, and serum albumin levels to determine whether B2M or CysC should be used to investigate a creatinine eGFR in the hyperfiltration or the 40-60 ml/min/1.73m2 range. Elderly individuals should have serum B2M measured as this biomarker is not affected by age^39^. On the other hand, patients with hypoalbuminemia should have CysC measured for the confirmatory testing.

Our study had also some limitations. The number of study participants was relatively small, as the study procedures were interrupted by the COVID19 pandemic. However, the study sample was diverse with significant racial and ethnical representation reflecting the target population. A significant proportion of our cohort had elevated hs-CRP levels; a sign of being in an inflammatory state. This status may have influenced the eGFR results we obtained from the various equations, especially the ones based on serum CysC and serum B2M levels that are influenced by chronic inflammation. Finally, the COMPASS study did not measure GFR using gold standard measurements such as inulin excretion rate, so our data are limited to evaluating discordances against three, imperfect biomarkers and prediction equations.

Considering the controversy of racial adjustments for eGFR estimation, we feel that B2M formula should be strongly considered as a biomarker of kidney function.

### Conclusions

Accurate estimation of GFR with common laboratory markers to appropriately identify CKD, predict prognosis and facilitate accurate drug dosing remains challenging. The diversity of the patient population needs to be taken into consideration when developing a new eGFR formula. Based on our predominant American Indian or Hispanic study population, we propose the use of B2M and CysC as alternative kidney function biomarkers when confirmation of a creatinine eGFR is needed in the hyperfiltration or the CKD stage 3a range.

### Support

The COMPASS study was supported by an unconditional grant from Dialysis Clinic, Inc. (DCI #C-3765 to C.A.). This project was supported by an award from the National Center for Advancing Translational Sciences, National Institutes of Health under grant number UL1TR001449.

## Authors’ Contributions

Research idea and study design: MMB, MR, CA; data acquisition: MMB, MR, CA; data analysis/interpretation: MMB, MR, CA; statistical analysis: MMB, MR, CA; supervision or mentorship: MR, CA. Each author contributed important intellectual content during manuscript drafting or revision and accepts accountability for the overall work by ensuring that questions pertaining to the accuracy or integrity of any portion of the work are appropriately investigated and resolved.

## Data Availability

All data produced in the present study are available upon reasonable request to the authors

## Acknowledgements

We also would like to thank UNM CTSC for research coordination.

