## Supplement Table 1 for "Comparison of Glomerular Filtration Rate Equations in a Rural New Mexico Cohort: Results from the COMPASS Study"

Female (S_cr_ ≤0.7), eGFR = 142 × (S_cr_/0.7)^-0.241^ × (0.993)^Age^ x 1.012

(S_cr_ >0.7), eGFR = 142 × (S_cr_ /0.7)^-1.209^ × (0.993)^Age^	x 1.012

Male	(S_cr_ ≤0.9), eGFR = 142 × (S_cr_/0.9)^-0.302^ × (0.993)^Age^

(S_cr_ >0.9), eGFR = 142 × (S_cr_/0.9)^-1.209^ × (0.993)^Age^

2) **2012 CKD-EPI Cystatin C equation**

Female (S_cr_ ≤0.8), eGFR = 133 x (S_cys_/0.8)^-0.499^ x 0.996^Age^ x 0.932

(S_cr_ >0.8), eGFR = 133 x (S_cys_/0.8)^-1.328^ x 0.996^Age^ x 0.932

Male	(S_cr_ ≤0.8), eGFR = 133 x (S_cys_/0.8)^-0.499^ x 0.996^Age^ X 1

(S_cr_ ≤0.8), eGFR = 133 x (S_cys_/0.8)^-1.328^ x 0.996^Age^ X 1

3) **2021 CKD-EPI Cystatin-Creatinine equation**

Female (S_cr_ ≤0.7 and S_cys_ ≤0.8), eGFR = 135 × (S_cr_/0.7)^-0.219^ x (S_cys_/0.8)^-0.323^ × (0.993)^Age^ x 0.963

(S_cr_ ≤0.7 and S_cys_ >0.8), eGFR = 135 × (S_cr_/0.7)^-0.219^ x (S_cys_/0.8)^-0.778^ × (0.993)^Age^ x 0.963

(S_cr_ >0.7 and S_cys_ ≤0.8), eGFR = 135 × (Scr/0.7)^-0.544^ x (S_cys_/0.8)^-0.323^ × (0.993)^Age^ x 0.963

(S_cr_ >0.7 and S_cys_ >0.8), eGFR = 135 × (Scr/0.7)^-0.544^ x (S_cys_/0.8)^-0.778^ × (0.993)^Age^ x 0.963

Male	(S_cr_ ≤0.9 and S_cys_ ≤0.8), eGFR = 135 × (S_cr_/0.9)^-0.144^ x (S_cys_/0.8)^-0.323^ × (0.993)^Age^

(S_cr_ ≤0.9 and S_cys_ >0.8), eGFR = 135 × (S_cr_/0.9)^-0.144^ x (S_cys_/0.8)^-0.778^ × (0.993)^Age^

(S_cr_ >0.9 and S_cys_ ≤0.8), eGFR = 135 × (S_cr_/0.9)^-0.544^ × (S_cys_/0.8)^-0.323^ x (0.993)^Age^

(S_cr_ >0.9 and S_cys_ >0.8), eGFR = 135 × (S_cr_/0.9)^-0.544^ × (S_cys_/0.8)^-0.778^ x (0.993)^Age^

4) **CKD-EPI Beta-2 Microglobulin equation****^12^**

eGFR$\beta2M$= 133 x β_2_M^-0.854^
